# Brains Under Stress: Unravelling the Effects of the COVID-19 Pandemic on Brain Ageing

**DOI:** 10.1101/2024.07.22.24310790

**Authors:** Ali-Reza Mohammadi-Nejad, Martin Craig, Eleanor Cox, Xin Chen, R. Gisli Jenkins, Susan Francis, Stamatios N. Sotiropoulos, Dorothee P. Auer

## Abstract

The impact of SARS-CoV-2 and the COVID-19 pandemic on cognitive and mental health is recognised, yet specific effects on brain health remain understudied. We investigated the pandemic’s impact on brain ageing using longitudinal neuroimaging data from the UK Biobank. Brain age prediction models were trained from hundreds of multi-modal imaging features using a cohort of 15,334 healthy participants. These models were then applied to an independent cohort of 1,336 participants with two MRI scans: either both collected before the pandemic (“Control” groups), or one before and one after the pandemic onset (“Pandemic” group). Our findings reveal that, even with initially matched brain age gaps (predicted brain age vs. chronological age), the pandemic significantly accelerated brain ageing. The “Pandemic” group showed on average 11-month higher deviation of brain age gap at the second time point compared with controls. Accelerated brain ageing was more pronounced in males and those from deprived socio-demographic backgrounds, with average increases of 3.3 and 7 months, respectively. These deviations existed regardless of SARS-CoV-2 infection. However, accelerated brain ageing correlated with reduced cognitive performance only in COVID-infected participants. Our study highlights the pandemic’s significant impact on brain health, beyond direct infection effects, emphasising the need to consider broader social and health inequalities.

## Introduction

Apart from the well-documented respiratory and systemic manifestations of SARS-CoV-2, compelling evidence highlights its neurotropic nature, showing high rates of persistent respiratory symptoms, fatigue, depression, post-traumatic stress disorder, and cognitive impairment in COVID-19 survivors^1^. Emerging research has revealed potential associations between COVID-19, cognitive decline, brain changes^2^, and the molecular signatures of brain ageing^3^. Significant psychological distress and mental health issues were also reported during the early pandemic phases, especially among younger and vulnerable individuals^4^. Conversely, recent reviews suggest variable reductions in mental health service use^5^ and, across 134 cohort studies, no overall rise in mental health conditions was found in the general population, with minimal increase in depression symptoms and small negative effects in women^6^. Understanding the pandemic’s effects on brain health, considering infection status and socio-demographic factors, is crucial for addressing its long-term health consequences and broader public health implications.

The neuroinvasion of SARS-CoV-2 is well established^7^, with virus persistence shown up to 230 days post-infection^8^. Central nervous system manifestations are attributed to neuroinvasion, vascular damage, and immune responses^9^. Recent studies suggest COVID-19 may worsen neurodegenerative processes or contribute to age-related cognitive impairments^10^. Longitudinal assessments indicate higher cognitive decline risk among COVID-19 survivors compared with controls^11^. Serial brain MRI showed reductions in grey matter (GM) thickness and white matter (WM) integrity, possibly due to neurodegeneration, neuroinflammation, or sensory deprivation^2^. Beyond brain infection or systemic effects, the COVID-19 pandemic may have independently impacted brain ageing due to psychosocial stressors, social disruptions, and lifestyle changes, as evidenced in adolescents^12^.

While indirect evidence suggests COVID-19 infection may accelerate brain ageing and neurodegeneration, comprehensive studies on the broader pandemic’s impact on brain age are lacking, which we explore here. Advanced neuroimaging and machine learning techniques have enabled powerful brain age prediction models, estimating deviations from typical ageing trajectories as the brain age gap (BAG=difference between estimated brain age and chronological age). Seminal works^13–15^ laid the foundation for these models, refined with large-scale datasets, multi-modal imaging^16,17^, extensions to multi-organ age predictions^18^, and proven associations with mortality. Utilising these methodologies, we estimate brain age and investigate the impact of COVID-19 and the pandemic on brain age using longitudinal neuroimaging data.

We hypothesise that COVID-19 infection and the pandemic accelerated brain ageing. To test this, we utilised serial neuroimaging data from the UK Biobank (UKBB) study^19^. We trained a model using multi-modal imaging-derived phenotypes (IDPs) to predict individual BAG. The trained model was then applied to unseen participants with two brain scans, one before and one after the pandemic (“Pandemic” group) or both scans before the pandemic (“Control” group). We further assessed the impact of COVID-19 infection within the “Pandemic” group and explored putative moderating factors on brain ageing, such as sex and deprivation indices, and the interrelation with cognitive decline.

## Results

A brain age prediction model^16^ (Fig. 1a) was trained on MRI scans collected pre-March 2020 from 15,334 healthy middle-aged and older UKBB^19^ participants (‘training set’: 8,407 female; age [mean±SD]: 62.6±7.6 years). To minimise potential confounds in brain age estimation caused by disease effects and comorbidities, only healthy participants, defined by the absence of chronic disorders before their scans (see Suppl. Table S4 as in ^18,20^), were included for both training and unseen cohorts. Hundreds of multi-modal IDPs were extracted^21^ and used as regressors in the model after PCA-based dimensionality reduction (Fig. 1b). To improve accuracy, separate models were created based on GM and WM features for males and females^18^. These models were then applied to our unseen study cohort with two MRIs, comprising 1,336 healthy participants (770 female; age: 59.7±6.2 years –Suppl. Fig. S1). This cohort included the “Pandemic” group (G1: *N*=404, 247 female) with one brain scan before and one after the pandemic, and the “Control” group (G2: *N*=932, 523 female) with both scans before the pandemic. The groups were adjusted to be matched for age, sex, and other health markers (see Suppl. Table S1), with a mean time interval between scans of ∼27 months for all groups (Suppl. Fig. S1c). BAG was estimated at both time points, and the rate of change in BAG was calculated and normalised for the time interval as R_BAG_=(ΔBAG/ΔT).

**Fig. 1.**
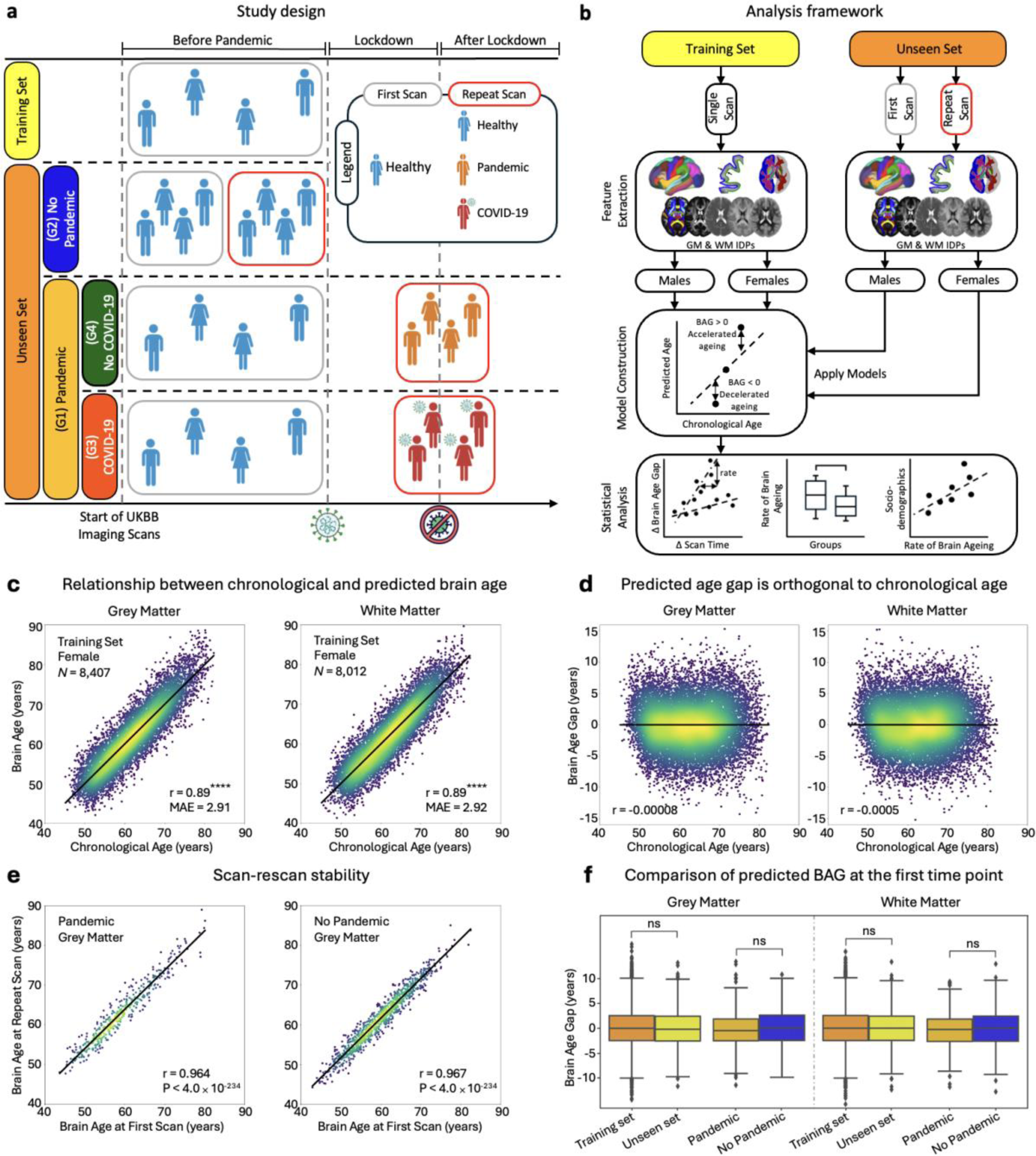
Study design, analysis framework, and accuracy assessment of brain age prediction models. a) The training set, consisting of healthy participants (free from any major chronic disease) with a single brain scan, is used to train a brain age prediction model with 20-fold cross-validation. This model is applied to the unseen set participants which includes the “Pandemic” group (G1), and the “No Pandemic” group (G2). The “Pandemic” group is subdivided into “Pandemic–COVID-19” (G3), and “Pandemic–No-COVID-19” (G4). b) The analysis framework starts with the extraction of imaging-derived phenotypes (IDPs) from grey matter (GM) and white matter (WM) tissues across different scan times. Separate predictive models are trained for different brain tissue types and sexes, applied to estimate the brain age gap at various time points. Statistical analyses are performed to compare groups and investigate the effects of COVID-19 and the pandemic using longitudinal data. c) Scatter plots show the relationship between chronological age (x-axis) and predicted brain age (y-axis) for GM and WM models in females (males in Suppl. Fig. S2). The diagonal line indicates where predicted age equals chronological age. ‘*N*’ is the number of subjects used for training. Accuracy is evaluated using Pearson correlation coefficient (r) and mean absolute error (MAE), averaged over 100 repetitions of 20-fold cross-validation. Asterisks (****) indicate p≤1.0e-04. d) This plot illustrates the relationship between chronological age and predicted brain age gap for GM and WM models, combining results for both sexes. The black regression line indicates deviations from perfect prediction, indicating zero age-related bias in the predicted age across the studied cohorts. e) The stability of the predictive model across two scans for participants in the “Pandemic” and “No-Pandemic” groups is evaluated, revealing a significant association between predicted brain ages from both scans (p<4e-234). The black regression line highlights deviations from perfect prediction. f) Boxplots compare predicted age gap distributions between the training set and unseen (first scan) set, and between the “Pandemic” and “No-Pandemic” groups for GM (left) and WM (right) models. The y-axis represents the predicted brain age gap in years. ‘ns’ indicates non-significance, showing similar mean predicted age gaps between unseen and training sets (GM: p=0.44, WM: p=0.99) and between “Pandemic” and “No-Pandemic” groups (GM: p=0.23, WM: p=0.28). Each data point in scatter plots represents an individual subject.

### Performance of brain age prediction models

Scatter plots in **Fig. 1**c (Suppl. Fig. S2) depict the relationship between chronological and predicted brain age for each brain tissue type and sex. We employed an unbiased estimation approach for brain age^16^, ensuring BAG is orthogonal to chronological age. All models demonstrated relatively similar prediction accuracy, with Pearson’s r ranging from 0.905 (WM female model, p<0.0001, 95%CI=0.901– 0.909; Mean Absolute Error (MAE)=2.90 years) to 0.894 (WM male model, p<0.0001, 95%CI=0.890– 0.899; MAE=3.09 years), indicating that neuroimaging features captured a large proportion of the chronological age variance, consistent with previous methodologically rigorous studies^18,22^.

We further confirmed that our model’s estimated brain age was unbiased towards the group mean^23^, and that participants’ age distribution was Gaussian^16^. **Fig. 1**d shows that there was also no significant correlation between the estimated BAG and chronological age when applying the trained model to unseen data (r<0.001)^†^. As expected, we found very high correlations between predicted brain ages of participants at the two time points (**Fig. 1**e, r>0.96), demonstrating high scan-rescan model reproducibility, further supporting that estimated brain ages reflect stable individual brain properties.

**Fig. 1**f shows the model’s generalisability, as we found no differences in mean predicted BAG between the training and unseen cohort’s first MRI data (Mann-Whitney two-sample t-test, GM: p=0.44, WM: p=0.99). Importantly, the estimated BAG for the first scan for the “Pandemic” group was not significantly different from the corresponding BAG for the “Control” (”No-Pandemic“) group (GM: p=0.23, WM: p=0.28), confirming that our matching (Suppl. Table S1) effectively achieved comparable baseline BAGs.

### Accelerated Brain ageing is associated with the COVID-19 pandemic, regardless of infection

Although BAGs were not statistically different between the “Pandemic” and “Control” groups at the first time point, the pandemic’s effect on brain ageing became evident with the second scan. Fig. 2 presents the rates of change in BAG between the two scans (R_BAG_). The “Pandemic” group displayed significantly higher R_BAG_ compared with the Controls (GM=12.5 months; WM=15.3 months; p<0.001), indicating accelerated brain ageing.

**Fig. 2.**
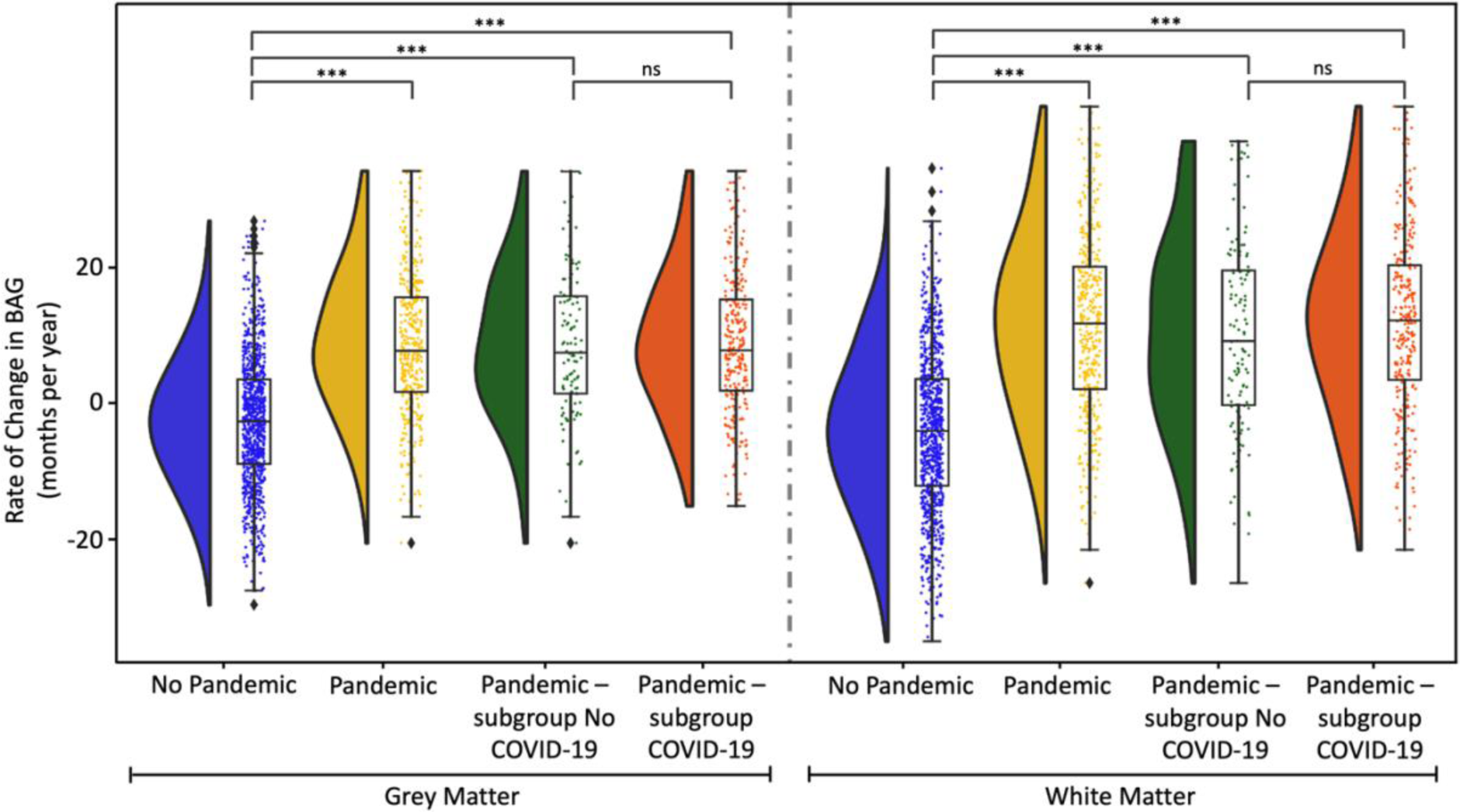
Effect of COVID-19 and the pandemic on brain ageing. This figure illustrates the distribution of the rate of change in brain age gap across different brain tissue models and subject groups. The left panel represents the Grey Matter model, and the right panel represents the White Matter model. Each subject group is depicted by different coloured violin plots: blue for the “No-Pandemic” group, orange for the “Pandemic” group, green for the “Pandemic–No-COVID-19” group, and red for the “Pandemic– COVID-19” group. The y-axis indicates the rate of change in brain age gap in months per year. Asterisks above the violin plot denote the results of two-sample t-tests comparing different groups: ‘ns’ indicates non-significance and *** indicates p-values in the range 0.0001< p ≤0.001.

To further investigate whether SARS-CoV-2 infection specifically influenced accelerated brain ageing, the “Pandemic” group was subdivided into: “Pandemic–COVID-19” (G3), with participants who had COVID-19 (121 participants–75 females, Suppl. Fig. S1e), and “Pandemic–No-COVID-19” (G4), with individuals without reported infection before the second scan (283 participants–172 females, Fig. 1a and Suppl. Fig. S1a-b). Notably, these subgroups were also adjusted to be matched to controls to ensure comparability (Suppl. Table S1). As illustrated in Fig. 2, both G3 and G4 had higher R_BAG_ values than “No-Pandemic” controls, with no significant difference between subgroups for either GM or WM models (Suppl. Fig. S3 shows brain age gap distributions at various time points across groups). This suggests increased positive brain age deviation (accelerated brain ageing) during the pandemic regardless of SARS-CoV-2 infection.

### Effects of age and sex on longitudinal brain ageing (rate of change in BAG)

While the estimated BAG was designed to be independent of chronological age for point estimates, biologically plausible longitudinal effects on the R_BAG_ cannot be excluded. We regressed R_BAG_ against chronological age (at first scan). Across all groups, a positive association between chronological age and accelerated brain ageing was observed (Suppl. Fig. S4). This association was stronger for the “Pandemic” group (Fig. 3a), showing older participants exhibited higher R_BAG_ in the “Pandemic” group compared with Controls.

**Fig. 3.**
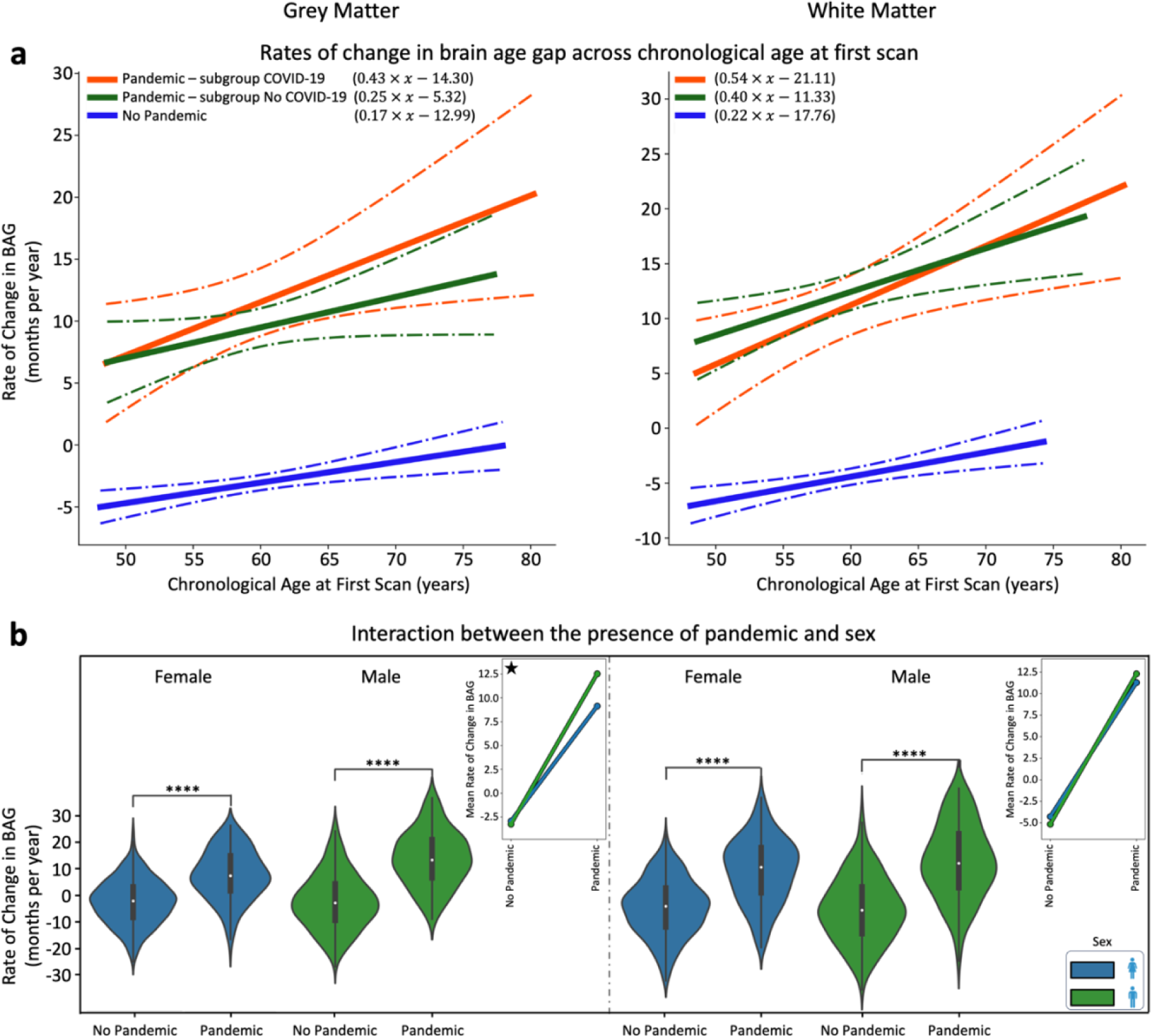
Impact of SARS-CoV-2 infection and the COVID-19 pandemic on accelerated brain ageing, and the role of age and sex in brain ageing during the pandemic. a) Evidence of accelerated brain ageing due to COVID-19 infection and the pandemic. Solid lines represent the best-fitted associations between the rate of change in brain age gap (y-axis) and the chronological age at first scan (x-axis) for “Pandemic–COVID-19”, “Pandemic–No-COVID-19”, and “No-Pandemic” groups. Dot-dashed curves depict the 95% confidence intervals around the best-fit lines. b) Violin plots display the distribution of the rate of change in brain age gap for “Pandemic” and “No-Pandemic” groups, stratified by sex. Asterisks indicate significance levels (**** signifies p-values ≤ 1.0e-04) from two-sample t-tests between the groups. Interaction plots on the right highlight distinct patterns in GM and WM between the “Pandemic” and “No-Pandemic” groups. Stars in the interaction plots indicate significant results, based on the interaction determined by the two-factor, two-level permutation test. GM model results are displayed on the left, and WM model results are shown on the right in both panels.

Specifically, each 1-year age increase at the first scan in Controls was associated with an approximate brain ageing acceleration of 5 days for GM (p=0.001) and 7 days for WM (p=0.0005) models. The “Pandemic” group demonstrated a twofold higher acceleration: brain ageing accelerated (10 and 14 days for GM (p=0.003) and WM (p=0.00006) respectively, Suppl. Fig. S4). A differential age-related acceleration of BAG was noted according to infection status. The strongest BAG augmentation by chronological age was observed in the COVID-19 infection subgroup (G3) with each 1-year increase in age at baseline showing 13 days of acceleration for GM (p=0.002) and 16 days for WM (p=0.006) (Fig. 3a), compared with 8 days for GM (p=0.007) and 12 days for WM (p=0.006) in those without infection (“Pandemic–No-COVID-19”).

The pandemic’s impact on accelerated brain ageing (higher R_BAG_ compared to Controls) was evident in both male and female participants (Fig. 3b; p<0.0001). We used two-factor, two-level permutation tests (5,000 permutations) to assess the interplay between the pandemic, sex, and their interactions on brain ageing. These tests confirmed the pandemic as significant factor for R_BAG_ (p=0.002 in both models—less than the 95%CI [0.0443–0.0564], calculated using the Wilson method^24^). In addition, sex was a significant factor in the GM model (p=0.018—less than the 95%CI [0.0443–0.0564]), but not in the WM model. Interestingly, a significant interaction (p=0.004—less than the 95%CI [0.0443–0.0564]) between sex and pandemic status was also found (for the GM model), indicating that the combination of the pandemic and being a male led to the highest R_BAG_ increases (33% more in males vs. females). The interaction plots (Fig. 3b) demonstrate divergence between males and females when comparing the “No-Pandemic” with the “Pandemic” group, highlighting the interaction between sex and the pandemic on GM-related brain ageing.

### Increased brain age gap rate during pandemic in deprived areas

Besides age and sex, socio-demographic factors can influence brain health and resilience to the pandemic. The effects of deprivation indices or drivers of poor brain health—such as health, employment, education, housing, and income—on brain ageing were examined.

The month-based clocks in **Fig. 4**a illustrate the extent of accelerated brain ageing among participants varying deprivation levels, highlighting changes from before to during the pandemic. The largest increases were seen in participants with different health scores (despite all participants being free from major chronic health conditions), showing a difference of about 5 months and 16 days in the GM model. This suggests an average increase of 5.5 months in R_BAG_ between participants with low vs. high health scores following exposure to the pandemic. Similarly, substantial changes were noted for low vs. high employment indices (5 months and 6 days increase), and low vs. high income levels (1 month and 29 days) in the GM model. The WM model showed significant R_BAG_ changes for low health index (7 months and 23 days increase), low employment index (6 months and 28 days), low education (4 months and 19 days), and low income (3 months and 8 days).

**Fig. 4.**
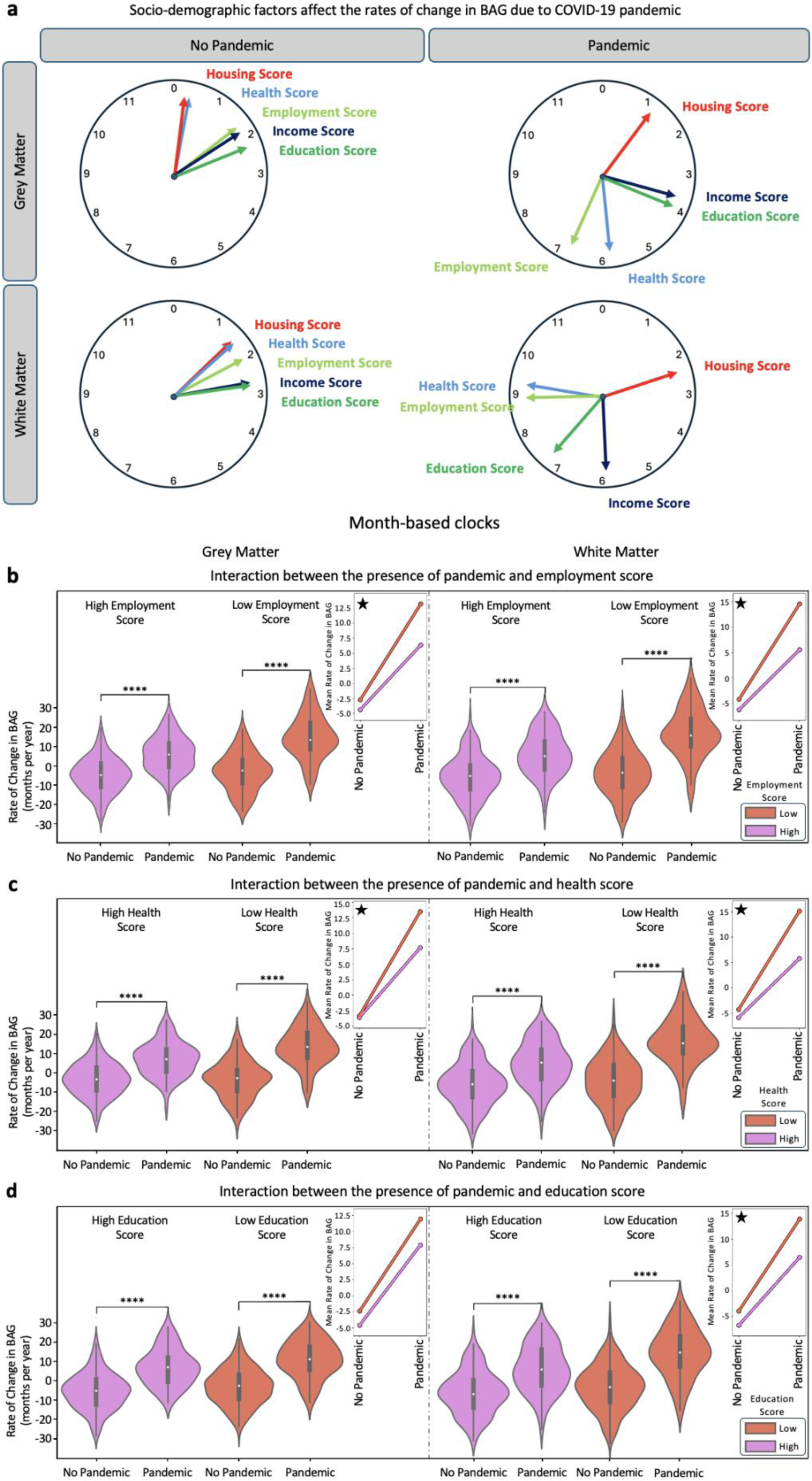
Influence of socio-demographic factors on brain ageing during the COVID-19 pandemic. a) The effects of socio-demographic factors, represented by indices of deprivation, on brain ageing in participants grouped by pandemic status. Each clock represents the difference in the mean rate of change in BAG between subjects with low and high levels of specific socio-demographic factors. The clocks are presented separately for GM and WM models, with one set depicting participants in the “No-Pandemic” group and another for participants in the “Pandemic” group. The socio-demographic factors studied include housing score, health score, employment score, income score, and education score. b-d) Violin plots display the distribution of the rate of change in BAG for the “Pandemic” and “No-Pandemic” groups in relation to (b) low (purple) vs. high (red) employment scores, (c) low (purple) vs. high (red) health scores, and (d) low (purple) vs. high (red) education scores. Each plot has two panels: the left side represents the GM model results, and the right side shows the WM model results. Asterisks indicate significance levels (**** signifies p-values ≤ 1.0e-04) calculated between the “Pandemic” and “No-Pandemic” groups using two-sample t-tests. Small plots on the right side of each panel depict interaction plots, suggesting the presence of interaction effects. These plots visualise how the mean rate of change in BAG deviates between the “No-Pandemic” and “Pandemic” groups in both GM and WM models. Stars in the interaction plots indicate significant results based on the two-factor, two-level permutation test, highlighting the interaction between the two factors.

Further analysis revealed significant differences (p<0.0001) in brain ageing patterns between the “Pandemic” and “No-Pandemic” groups across the deprivation indices (**Fig. 4**b-d). Generally, the increase in R_BAG_ between the “Pandemic” and Control groups was higher for participants with high deprivation scores (low health, low education, and low employment) compared to those with low deprivation scores (high health, high education, and high employment). This was true for both GM and WM models, indicating potential interactions between the pandemic’s effects and deprivation on brain ageing differences.

To further explore such interactions, we conducted non-parametric two-factor, two-level permutation tests. These tests confirmed the pandemic significantly drove the differences in predicted R_BAG_ between “Pandemic” and Control groups. Several deprivation indices also influenced differences between low and high deprivation, including employment (GM: p=0.0002; WM: p=0.0002), health (GM: p=0.0006; WM: p=0.0002), education (GM: p=0.0004; WM: p=0.0002), and income score levels (GM: p=0.0026; WM: p=0.0002) (all below 95%CI [0.0443–0.0564]). Housing scores were not significant in either model.

Significant interactions between pandemic status and deprivation factors were also found (<95%CI [0.0443–0.0564]). Specifically, interactions between pandemic status and employment (GM: p=0.0056; WM: p=0.0006), health (GM: p=0.0016; WM: p=0.0002), and education scores (WM: p=0.0134) on brain ageing were found to be significant. Fig. 4b-d depict interaction plots comparing distinct patterns in GM and WM models between the “Pandemic” and “No-Pandemic” groups, highlighting socio-demographic factors’ role in brain ageing during the pandemic. As sex significantly interacted with pandemic status only in the GM model (Fig. 3b), we also analysed the interplay of each deprivation index and pandemic status separately for female and male participants. Results showed that even in sex-specific models, all previous findings and interactions between pandemic and deprivation remained significant (Suppl. Fig. S5).

### Cognitive performance, accelerated brain ageing, and COVID-19 exposure

To assess the impacts of COVID-19 and the pandemic on cognitive performance related to longitudinal brain ageing, we analysed performance changes over time among individuals who completed cognitive tests at both scans. This analysis included three groups: “No-Pandemic”, “Pandemic–COVID-19”, and “Pandemic–No-COVID-19”, focusing on the top 10 cognitive tests related to dementia risk within the UKBB^2^.

Among these groups, the “Pandemic–COVID-19” group showed a significantly (**Fig. 5**, insets) higher decline in performance from baseline to follow-up only for one cognitive test (trail making test—TMT). Specifically, there were significantly larger increases in completion time for both TMT-A and TMT-B compared with the Control and “Pandemic–No-COVID-19” groups (**Fig. 5**). This indicates a notable decline in cognitive function specifically in those who had contracted COVID-19.

**Fig. 5.**
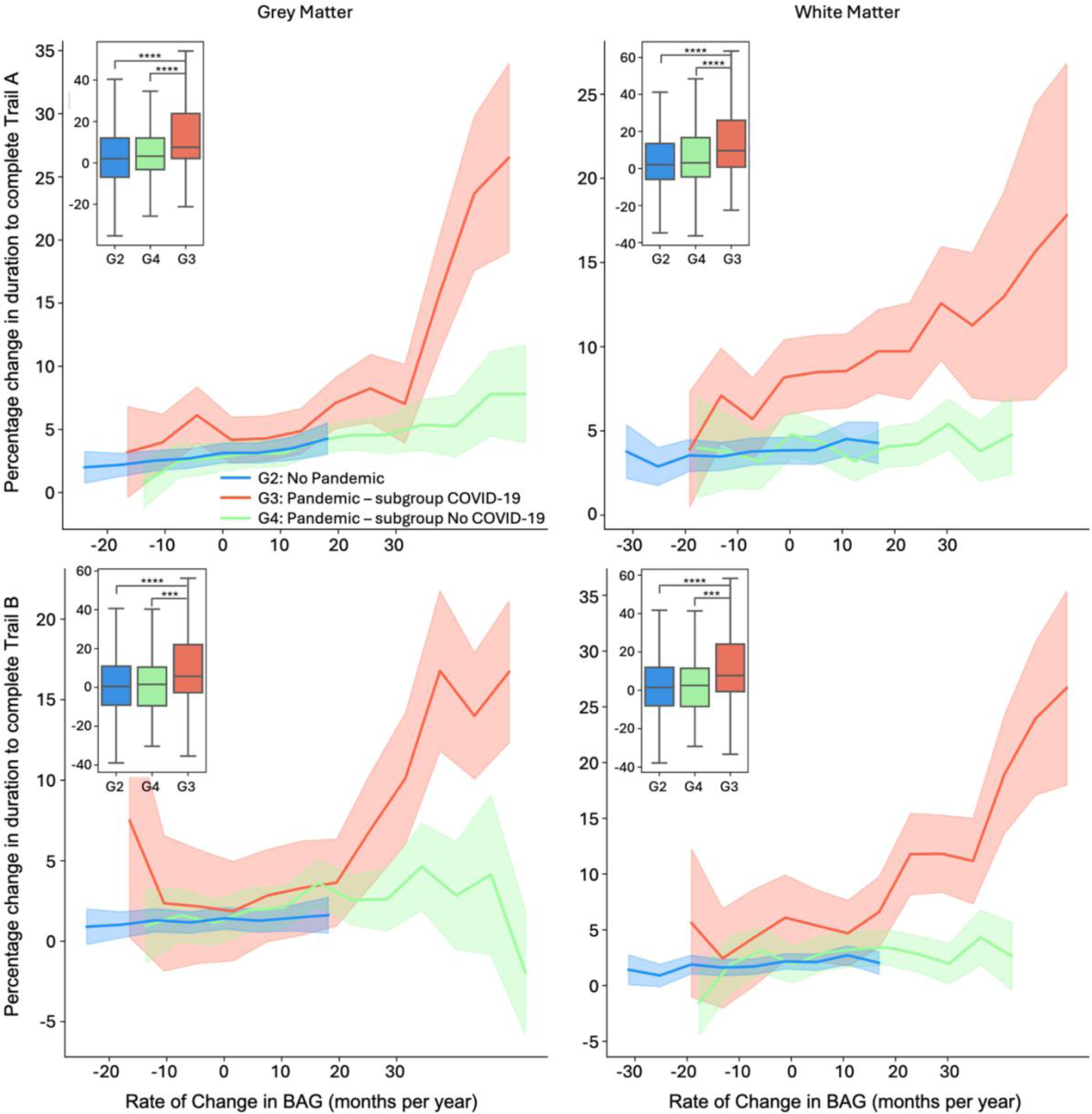
Impact of COVID-19 on cognitive performance across rates of change in brain age gap. The figure illustrates the percentage change in completing the (top) TMT-A, and (bottom) TMT-B over two imaging scans across various rates of change in brain age gap for participants in the “Pandemic–COVID-19” (red), “Pandemic–No-COVID-19” (green), and “No-Pandemic” (blue) groups, depicted using both GM (left) and WM (right) models. A three-year sliding window was used to create these curves. The standard error values are depicted in light blue (for the “No-Pandemic” group), light green (for the “Pandemic–No-COVID-19” group), and light red (for the “Pandemic–COVID-19” group). The boxplots in the top left of each plot show the distribution of percentage change in completing the TMT over time without a sliding window. Participants with COVID-19 (G4) had a greater percentage decline in performance (longer completion time) compared with the Control group (G2), with p-values of 1.3e-7 (Trail A–GM), 5.4e-7 (Trail A–WM), 6.8e-5 (Trail B–GM), and 6.8e-6 (Trail B–WM) against a Bonferroni threshold of 8.33e-4. Significant differences were also noted between COVID-19 infected (G4) and non-infected (G3) Pandemic participants (p(Trail A–GM)=3.6e-5, p(Trail A–WM)=4.8e-5, p(Trail B– GM)=7.4e-4, p(Trail B–WM)=1.1e-4). Asterisks indicate the significance levels (*** signifies p-values ≤ 1.0e-03 and **** signifies p-values ≤ 1.0e-04) calculated between different groups using two-sample t-tests.

Further analysis revealed differences in the association between R_BAG_ and TMT-A performance across the groups and models, using full and partial correlations excluding chronological age. A small but significant positive linear association was noted between brain ageing changes and cognitive decline in both the “No-Pandemic” and “Pandemic-No-COVID-19” groups for both GM and WM models (Suppl. Fig. S6). In contrast, the “Pandemic–COVID-19” group showed a more pronounced and non-linear decline in cognitive performance with higher R_BAG_, suggesting a differential threshold effect between GM and WM models. These findings suggest that while BAG increase during the pandemic was independent of COVID-19 infection, it was only associated with decline in one cognitive test (TMT), and only in those with recorded COVID-19 (G3).

## Discussion

Using longitudinal neuroimaging data from the UKBB, we estimated individual brain age, and its change rate compared to chronological ageing in two matched cohorts: one scanned before and during the COVID-19 pandemic, and the other scanned twice before the pandemic. We found that the COVID-19 pandemic was detrimental to brain health and induced accelerated brain ageing for GM and WM derived models, regardless of SARS-CoV-2 infection. Accelerated brain ageing during the pandemic was more pronounced in older individuals and males based on the GM model, and in those from deprived backgrounds for both models. Cognitive performance, particularly in flexibility and processing speed tasks, declined significantly in COVID-19 infected individuals, correlating with accelerated GM ageing. Conversely, participants who experienced the pandemic without reported infection had similar age-related declines as controls, demonstrating that pandemic-related accelerated brain ageing alone was insufficient to lead to cognitive decline.

These findings provide novel insight into how the COVID-19 pandemic affected brain health, demonstrating that the general pandemic effects alone, without infection, exerted a substantial detrimental effect on brain health augmented by bio-social factors (age, health, and social inequalities) in a healthy middle-aged to older population. Notably, the extent of accelerated brain ageing over a matched pre-pandemic control group, observed in grey and white matter, was similar in both non-infected and infected sub-cohorts. This highlights the major role of pandemic-related stressors such as anxiety, social isolation, and economic, and health insecurity on brain changes that may be sufficient to explain the observed accelerated brain ageing. In other words, our findings suggest that a full bio-psycho-social model is needed to understand the negative brain health effects of COVID-19 infection during a pandemic, which previous research has not accounted for^2^.

Our findings and interpretation align well with reports of increased internalising symptoms, reduced cortical thickness, and accelerated brain ageing in adolescents during the pandemic^12^. Nevertheless, in the age group we studied, advanced brain ageing directly indicates poor brain health, thus avoiding complexities of adolescent brain maturation. The most plausible explanation for the observed accelerated brain ageing is chronic stress and social isolation, consistent with well-documented sequelae like neuroinflammation, structural and functional brain changes in preclinical models^25–29^. Previous studies in humans confirm that social isolation and perceived loneliness contribute to structural and functional brain changes^30,31^ that are expected to drive the observed accelerated brain ageing.

We further explored how pandemic-related stressors like social isolation, economic hardships, and healthcare disruptions interact with pre-existing health disparities and age to affect brain health^37,38^. These stressors disproportionately affect vulnerable populations, worsening mental health and amplifying inequalities^39,40^. We report notable augmenting effects on the pandemic-related accelerated brain ageing for a range of deprivation factors, most pronounced for low employment, low education and low health scores. The observed interactions provide new and quantitative evidence for the differential effect of the COVID-19 on brain health across the population with substantial widening of the brain age gap in socially and economically disadvantaged groups^41,42^. It remains unclear whether the brain ageing effects may be at least partially be reversible, but the strong link to deprivation further emphasises the urgent need for policies addressing health and socio-economic inequalities, as the pandemic has exacerbated pre-existing disparities^43–45^.

Previous studies have documented SARS-CoV-2’s neural and vascular impacts, including inflammation and secondary systemic infection effects^46,47^. Our findings provide additional evidence of accelerated brain ageing in middle-aged to older participants with asymptomatic and mild-to-moderately affected COVID-19 infection without major comorbidities. This accelerated BAG effect was independent of infection status. Nevertheless, we show a complex and partially differential effect of old age. While the BAG model is by design independent of chronological age, BAG change was higher in older age in all groups including Controls, suggesting that age-related mechanisms contribute to the observed accelerated brain ageing^48^. This effect was strongest for the GM model for COVID-19-infected participants, which may offer an explanation for the observed differential effect on cognition. Cognitive decline is well-documented in ageing^49,50^, and we confirm faster cognitive decline in older appearing brains during the pandemic in all groups. However, we report a distinctly more pronounced age effect in COVID-infected participants with an apparent threshold suggesting a complex three-factor model of cognitive decline due to more pronounced accelerated brain ageing from pandemic-related stressors and additional infection-related factors in older people. This supports the concept of brain resilience loss leading to faster cognitive decline, consistent with existing neurodegeneration and dementia research^2,51–53^ and novel epigenetic models^54,55^.

It is conceivable that additional factors may have contributed to accelerated brain ageing during the pandemic ^2,35^in both infected and non-infected subgroups such as reduced physical activity, poorer diets, and increased alcohol consumption, all negatively impacting brain health^32–34^. Our study focused on cumulative, easier to interpret brain ageing effects limiting the ability to dissect region and modality specific features that may disentangle diverse pathomechanisms. Nevertheless, differences in our findings derived from GM and WM models highlight potential implications for understanding neurodegeneration and other brain health issues^2,36^. Further research should explore specific GM and WM features driving acceleration of brain ageing that may allow to disentangle tissue, regional and imaging marker-specific features that can be linked to neuro-glial-vascular mechanisms of brain ageing.

Our study has notable strengths and limitations. Employing BAG models provided an interpretable brain-wide health marker that was sensitive to disentangle contributory biopsychosocial factors exploiting the power of a longitudinal imaging-rich population study before and during the pandemic. We extended evidence on brain changes due to COVID-19 and socio-economic deprivation^2,45^. The careful subgroup comparison highlights that the main brain ‘cost’ of the pandemic was not due to infection itself, though causal inference cannot be claimed^56^. More research is necessary to establish causal relationships between deprivation factors and accelerated brain ageing, considering complex interactions. The study is further limited by access to only two time points, prohibiting assessment of potential reversibility. Longer follow-ups after the pandemic are needed to investigate persistent brain ageing effects and their long-term consequences beyond acute cognitive impacts in the infected subgroup. Furthermore, the time interval between scans differed between groups, with a wider spread in the Pandemic group compared with Controls. To mitigate this, we excluded Pandemic subjects to match the mean time interval to that of Controls (see Methods-Study Design). We found that the mean and reduced spread of time intervals used in our analyses maintained and slightly increased the R_BAG_ differences compared to results from repeating the analyses with the original unmatched Pandemic group. This suggests that with an ideal match of time intervals across groups, the same trends and differences (if not larger) should be observed.

In conclusion, the COVID-19 pandemic profoundly impacted brain health, shown as accelerated brain ageing, influenced by bio-psycho-social factors, especially social and health deprivation. Notably, the main effects were independent of infection status, except for interactions between COVID-19 infection, brain ageing, old age, and cognitive decline. Our findings highlight the need to address health and socio-economic inequalities in addition to lifestyle factors to mitigate accelerated brain ageing. Continued research and targeted policies are crucial to improve brain health outcomes in future public health crises.

## Methods

### Study Design and Neuroimaging Data

The UKBB imaging study provided multi-modal brain imaging data^19^ from over 42,677 participants (released in April 2023), aged 45 and older, collected at four sites with identical protocols, ensuring high-quality data. Before the COVID-19 pandemic, around 3,000 participants underwent a second scan as part of a longitudinal study. Starting in February 2021, an additional 2,000 participants were re-scanned to investigate the effects of SARS-CoV-2, bringing the total with repeat scans to about 5,000.

Re-imaging study participants met specific criteria: no incidental findings from initial scans, residence within a defined catchment area, and high-quality initial scans. SARS-CoV-2-positive participants were identified through diagnostic tests, primary care data, hospital records, or antibody tests. Controls were matched 1:1 to positive cases by sex, ethnicity, birth date, imaging site, and initial imaging date.

We excluded participants with chronic disorders before their scans (e.g., dementia, schizophrenia) to avoid bias in brain age predictions^18,20^ (Suppl. Table S4). Those with low-quality anatomical MRI data^21^ or unreliable brain IDPs were also removed. Technical outliers were defined as IDP values greater than or equal to five times the standard deviation from the cohort mean IDPs. Subjects with high levels of missing or unreliable IDPs in any session were excluded.

For training the brain age prediction model^16^, we used participants with one imaging session collected before March 2020 (**Fig. 1**a, *N* = 15,334; 8,407 female; age range: 45.1–82.4 years; mean ± SD: 62.6 ± 7.6 years). The unseen set included those with two scans (*N* = 1,336; 770 female; age range: 48.1–80.2 years; mean ± SD: 59.7 ± 6.2 years), divided into “Pandemic” (G1) and “No-Pandemic” (G2) groups. The “Pandemic” group (G1) included subjects with scans before and after the pandemic onset (*N* = 404; 247 female), further split into “Pandemic–COVID-19” (G3, *N* = 121; 75 female) and “Pandemic– No-COVID-19” (G4, *N* = 283; 172 female) (**Fig. 1**a). Groups were matched based on gender, age, BMI, alcohol intake, smoking, blood pressure, education, deprivation index, and general health metrics. (Suppl. Table S1, Suppl. Fig. S1).

The time interval between scans differed between groups, with a wider spread in the “Pandemic” group compared to Controls, due to the timings of the data acquired during the pandemic. Including all available participants would have resulted in a time interval of 39.0±18.6 months for the Pandemic group vs. 27.1±1.5 months for Controls. Instead, we excluded 255 subjects from the Pandemic group to achieve a time interval distribution of 26.8±5.6 months, reducing the spread (5.6 vs. 18.6 months) and matching the mean time interval (26.8 months vs 27 months) to Controls.

### Brain Age Modelling

We trained a multivariate regression model to assess brain ageing by regressing imaging-derived phenotypes (IDPs) against participants’ ages. This estimated an individual’s brain age *Y*_*B*_ and calculated the brain age gap (BAG), defined as *δ* = *Y*_*B*_ − *Y*, where *Y* is the chronological age. A positive δ (*δ* > 0) indicates an older-appearing brain, while a negative δ (*δ* < 0) indicates a younger-appearing brain. Age was modelled as a function of *M* imaging-derived phenotypes, *Y*_*B*_ = *f*(***X***), with ***X*** being a matrix of dimensions *N* × *M*, where *N* is the number of participants. We used a general linear regression method introduced by Smith et al.^16^ to ensure an unbiased *δ* orthogonal to chronological age.

Following the methodology of ^18^, we trained separate models for males and females, and for grey matter (GM) and white matter (WM), using IDPs from a healthy cohort free from chronic medical conditions (**Fig. 1**b). To reduce the dimensionality of the imaging features matrix, we applied singular value decomposition, retaining the top 50 eigen-subject-vectors that explain the most variance in ***X***^16^.

A 20-fold cross-validation process was used to train the model. In each iteration, a linear regression model was trained on 19 folds, and the fitted coefficients were applied to the held-out fold to predict brain age. During prediction, we de-confounded test set measures using the regressor’s weights from identified confounding variables in the training set, following the approach used by Miller et al.^19^. Notably, age-dependent confounds were not removed from the IDPs. To ensure robustness, this cross-validation process was repeated 100 times with random assignment to folds in each trial, affirming the reliability of our brain age estimation model.

Post-training, the age prediction models were applied to unseen data from G1 and G2 groups, for both females and males (Fig. 1a). Predictions were performed independently for initial (*t*_1_) and repeat scans (*t*_2_) of participants, allowing estimation of brain age gaps *δ*_*t*1_ and *δ*_*t*2_, respectively. The rate of change in BAG (R_BAG_) was then calculated as (*δ*_*t*2_ − *δ*_*t*1_)⁄*T*, where *T* is the chronological age difference between the two measurement time points^18,57^.

### Feature Selection for Brain Age Modelling

We selected IDPs to build predictive models for brain age estimation, focusing on both GM and WM regions (Fig. 1b). For the GM model, structural IDPs from T_1_-weighted MRI scans^19,21^ included the volume of subcortical structures, cortical/cerebellar regions, cortical surface area, cortical thickness, and GM/WM intensity contrast, totalling *M* = 1,422 IDPs. The WM model used IDPs from T_2_-weighted and diffusion MRI scans, assessing tissue complexity and integrity using diffusion tensor imaging (DTI) and neurite orientation dispersion and density imaging (NODDI) metrics, including fractional anisotropy (FA), mean diffusivity (MD), eigenvalue maps, and more, totalling 443 IDPs. CSF-related IDPs were excluded. Selected IDPs are listed in Suppl. Tables S2 and S3.

IDPs were de-confounded following^19,58^, considering 46 variables (Suppl. Tables S5), such as head size, sex, head motion, scanner table position, imaging centre, and scan-date-related drifts, excluding age-related confounds^59^.

### Interaction Effects against Socio-Demographic Factors

After calculating IDP-based brain age gaps and rates, we investigated interactions between brain ageing and socio-demographic factors using permutation-based inference with FSL PALM^60^.

We conducted a series of 2-way analyses (permutation-based ANOVA) to examine the rate of change in BAG between two time points. Factor 1 was the pandemic presence; Factor 2 included socio-demographic variables: sex, regional employment, health, education, housing, and income scores. The latter five factors are indicators of deprivation (detailed descriptions of these indices can be found in the Suppl. Material).

For each model tested, we assessed whether significant main effects existed—specifically, whether factor 1 (pandemic presence) or factor 2 (socio-demographic variable) had a discernible impact on brain ageing. Additionally, we explored interaction effects to determine if the combined influence of both factors produced a different impact on brain aging compared to their individual effects.

### Cognitive Scores

We selected the top 10 cognitive tests from the UKBB that have been associated with dementia risk^2^ (see Suppl. Table S6). To compare participants’ cognitive abilities across different groups, we calculated the percentage change in their cognitive scores between the two scans. This was done using the formula: *Percentage Change* = (*Score*_*t*2_ − *Score*_*t*1_) × 100⁄*Score*_*t*1_ where *Score*_*t*2_ and *Score*_*t*1_ represent the cognitive test results at the second and first time points, respectively.

## Data Availability

Scripts for estimating brain-age using imaging-derived features are available on GitHub https://github.com/SPMIC-UoN/deltab.

## Contributions

AM: data curation, methodology, formal analysis, writing – original draft, MC: methodology, writing – review & editing, EC: writing – review & editing, XC: writing – review & editing, RGJ: funding acquisition, writing – review & editing, SF: funding acquisition, writing – review & editing, SNS: methodology, resources, supervision, funding acquisition, conceptualisation, writing – original draft, DPA: methodology, supervision, conceptualisation, funding acquisition, writing – original draft

## Data sharing

All data used in this study are publicly available from the UK Biobank (www.ukbiobank.ac.uk). Our study was performed under UK Biobank Project 43822, PI: Sotiropoulos. Scripts for estimating brain-age using imaging-derived features are available on GitHub https://github.com/SPMIC-UoN/deltab.

## Acknowledgements

Data were provided by the UK Biobank under Project ID 43822 (PI: Sotiropoulos). This study was supported by the NIHR-funded Nottingham Biomedical Research Centre and the DEMISTIFI consortium – (Medical Research Council grants MR/V005324/1, MR/W014491/1). SNS acknowledges funding from the European Research Council (ERC Consolidator 101000969). The computations described in this paper were performed using the University of Nottingham’s Ada HPC service and the Precision Imaging Beacon Cluster, which provide High-Performance Computing service to the University’s research community.

## Footnotes

† For the remainder of this paper (including Fig. 1d), unless otherwise stated, we aggregated the predicted brain age gap for male and female models across different brain tissue types and participant groups separately.

