## Supplementary Materials for "Brains Under Stress: Unravelling the Effects of the COVID-19 Pandemic on Brain Ageing"

### Suppl. Materials

#### *Health Outcomes and Clinical Characterization*

To mitigate potential bias in brain age caused by disease effects, we excluded subjects who had any type of chronic disorders (such as dementia, parkinsonism, multiple sclerosis (MS), schizophrenia, depression, bipolar disorder, or cancer)[1,2] before their scans. The full list of 17 chronic diseases is in Table S4. These conditions have been identified using main and secondary ICD-10 codes (Data-Fields 41202, 41204, 41270, and 41271) and self-reports (Data-Fields 20001 and 20002) available in the UK Biobank dataset. For each individual, the recorded date of diagnosis has been compared across self-report (Data-Fields 20006 and 20008) and healthcare (Data-Fields 41262, 41280, and 41281) sources for each disease category, to determine whether the illness onset/diagnosis preceded or occurred after the first brain imaging scan.

#### *Multiple Indices of Deprivation*

In this study, we explored how the difference in the brain age gap relates to different measures of deprivation like education, employment, health, housing, and income scores. These scores come from a study of deprived areas in British local councils provided separately for England, Wales, and Scotland (Index of multiple deprivation Data-Fields: 26410, 26426, and 26427 – education score Data-Fields: 26414, 26421, and 26431 – employment score Data-Fields: 26412, 26419, and 26429 – health score Data-Fields: 26413, 26420, and 26430 – housing score Data-Fields: 26415, 26423, and 26432 – income score Data-Fields: 26411, 26418, and 26428).

These deprivation scores measure various aspects of hardship in small areas. This includes things like not having enough money, struggling to find a job, facing health issues, not getting a good education, having trouble finding housing or services, living in a poor environment, and being affected by crime. Each type of hardship is measured separately, using different indicators and people in an area might experience one or more of these kinds of deprivation. These measurements help us understand which areas are most affected by deprivation and what challenges people there face. So, we can create a ranking of areas based on their level of deprivation. Overall, these measures help to identify the most and least deprived areas in the country and provide information about the different challenges people face depending on where they live.

To simplify the scores, we categorised all the participants among these 3 groups into "high" and "low" levels. In each country, for education, employment, health, housing, and income scores, subjects scoring above the 70th percentile were classified as "high", while those below the 30th percentile were classified as "low". Similarly, for the index of multiple deprivations, subjects with indices above the 70th percentile were considered "low", and those below the 30th percentile were considered "high". Subjects with missing information were excluded from the analysis in all cases.

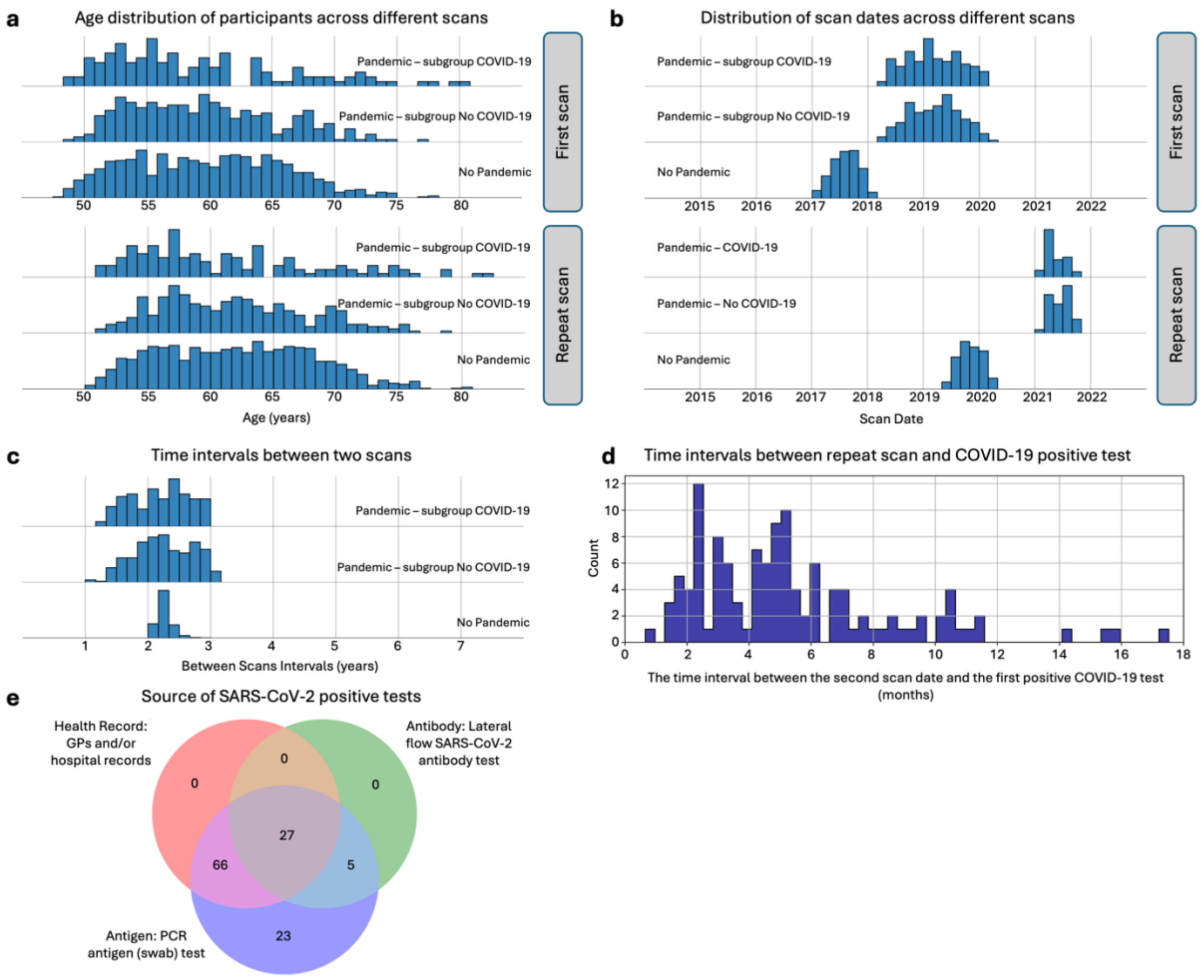


Fig. S1. The demographics of participants were categorised into “COVID-19”, “Lockdown”, and “No Pandemic” groups. a) The top section shows the age distribution of participants at Timepoint 1, while the bottom section displays the distribution at Timepoint 2 for each group. b) The distribution of assessment visit dates is depicted for both Timepoint 1 (top) and Timepoint 2 (bottom), showing the spread across different groups. c) The time intervals between two imaging scans are presented for each group, providing insights into the frequency of scans over time. d) Estimated dates of COVID-19 symptoms for the participants in the “COVID-19” group. e) The source of evidence for COVID-19 infection in the "COVID-19" group. Antibody – home-based lateral flow SARS-CoV-2 antibody test; Antigen – PCR antigen (swab) test; and Health records – from general practitioners (GPs) and/or hospital records.

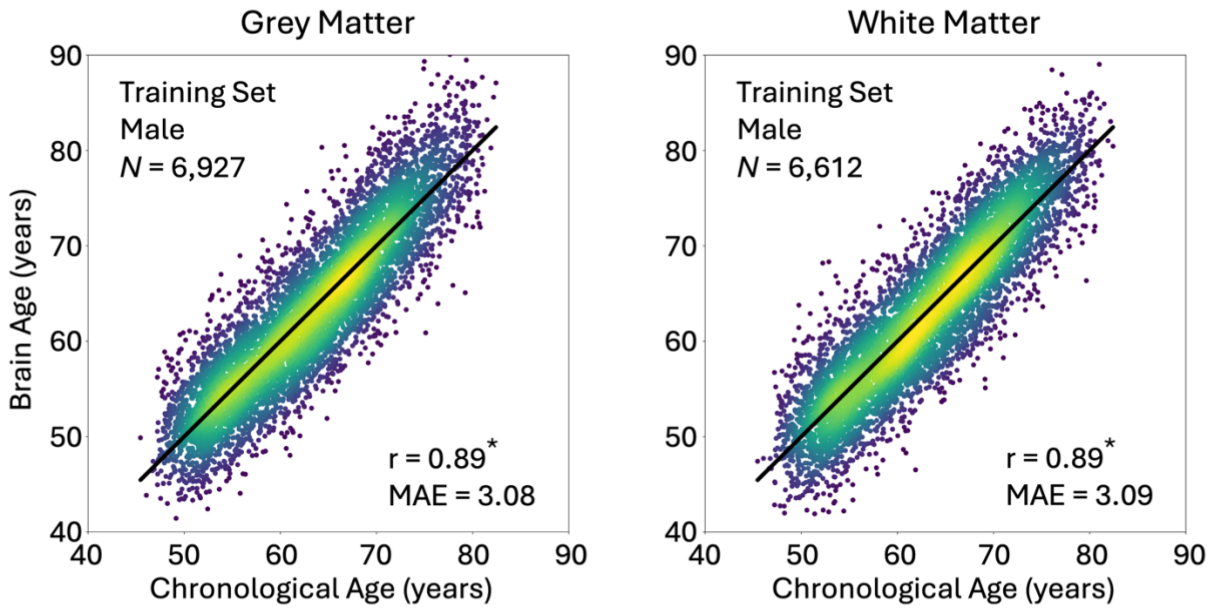


Fig. S2. The scatter plots show the relationship between chronological age (x-axis) and predicted brain age (y-axis) for GM and WM models, specifically for males.

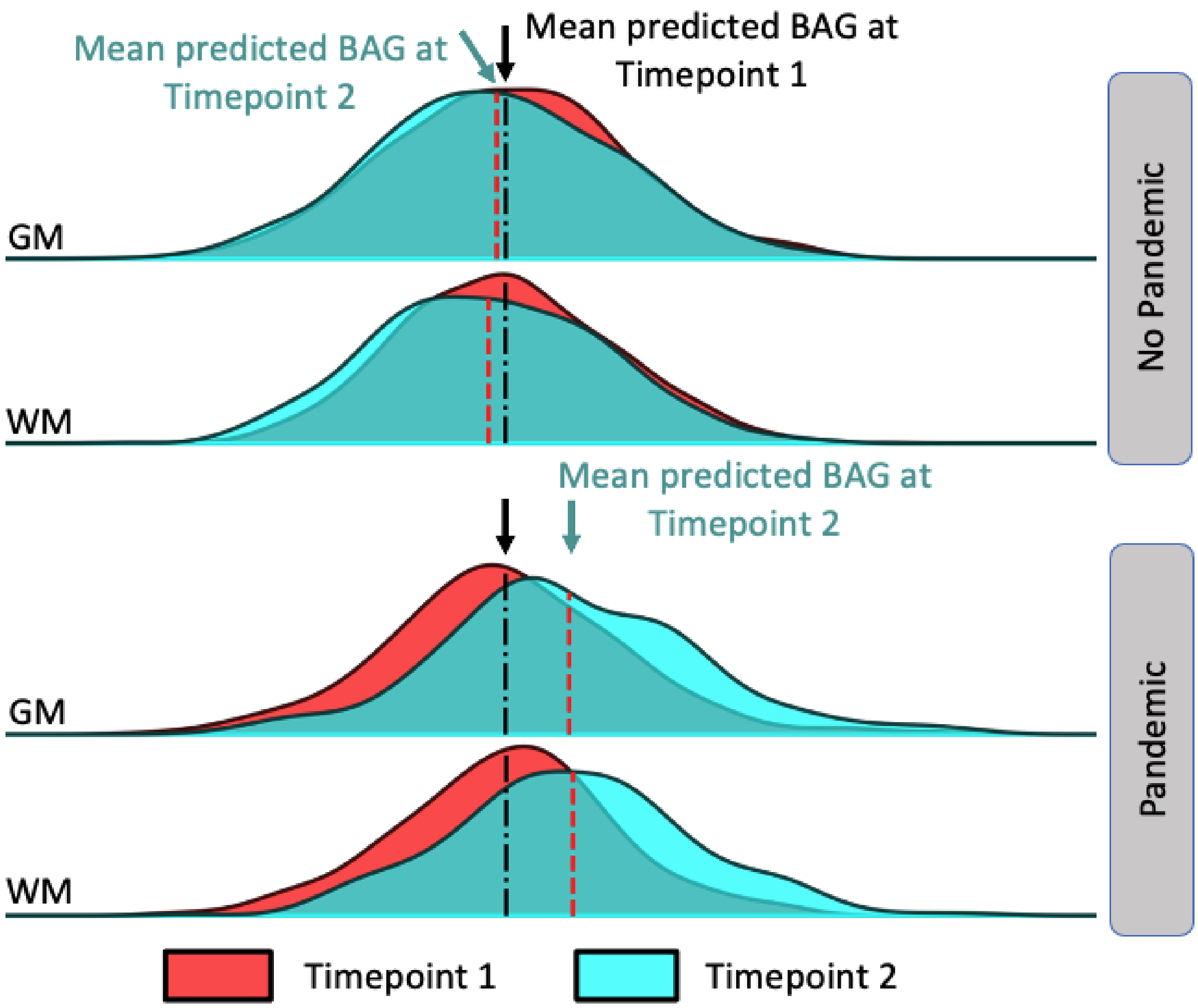


Fig. S3. The distribution of predicted brain age gaps at Timepoint 1 (red) and Timepoint 2 (cyan) in both GM and WM models. Black dash-dot lines represent the mean predicted age gap at Timepoint 1, while red dashed lines indicate the mean at Timepoint 2.

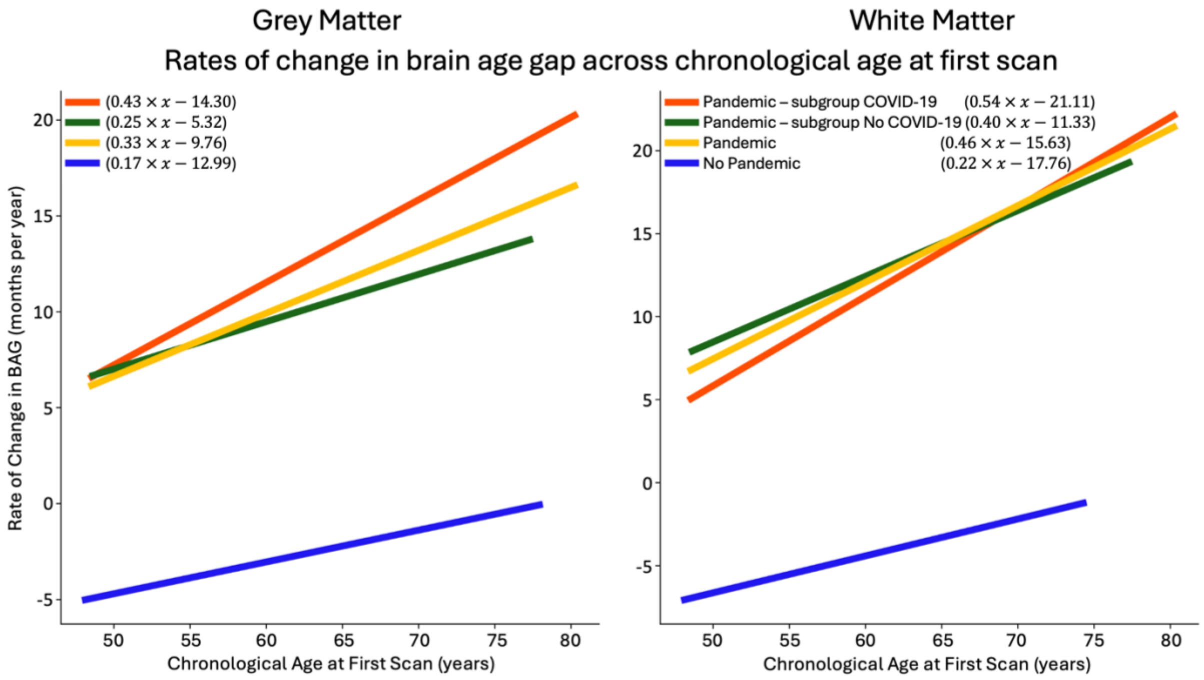


Fig. S4. Accelerated brain ageing observed in individuals with SARS-CoV-2 infection and those who experienced COVID-19 pandemic in both GM (left) and WM (right) models. The relationship between the rate of change in brain age gap (x-axis) and chronological age at first scan (y-axis). The solid lines represent the best-fitted associations between the x-axis and y-axis variables for different participant groups, while the dot-dashed curves depict the 95% confidence intervals around these best-fit lines.

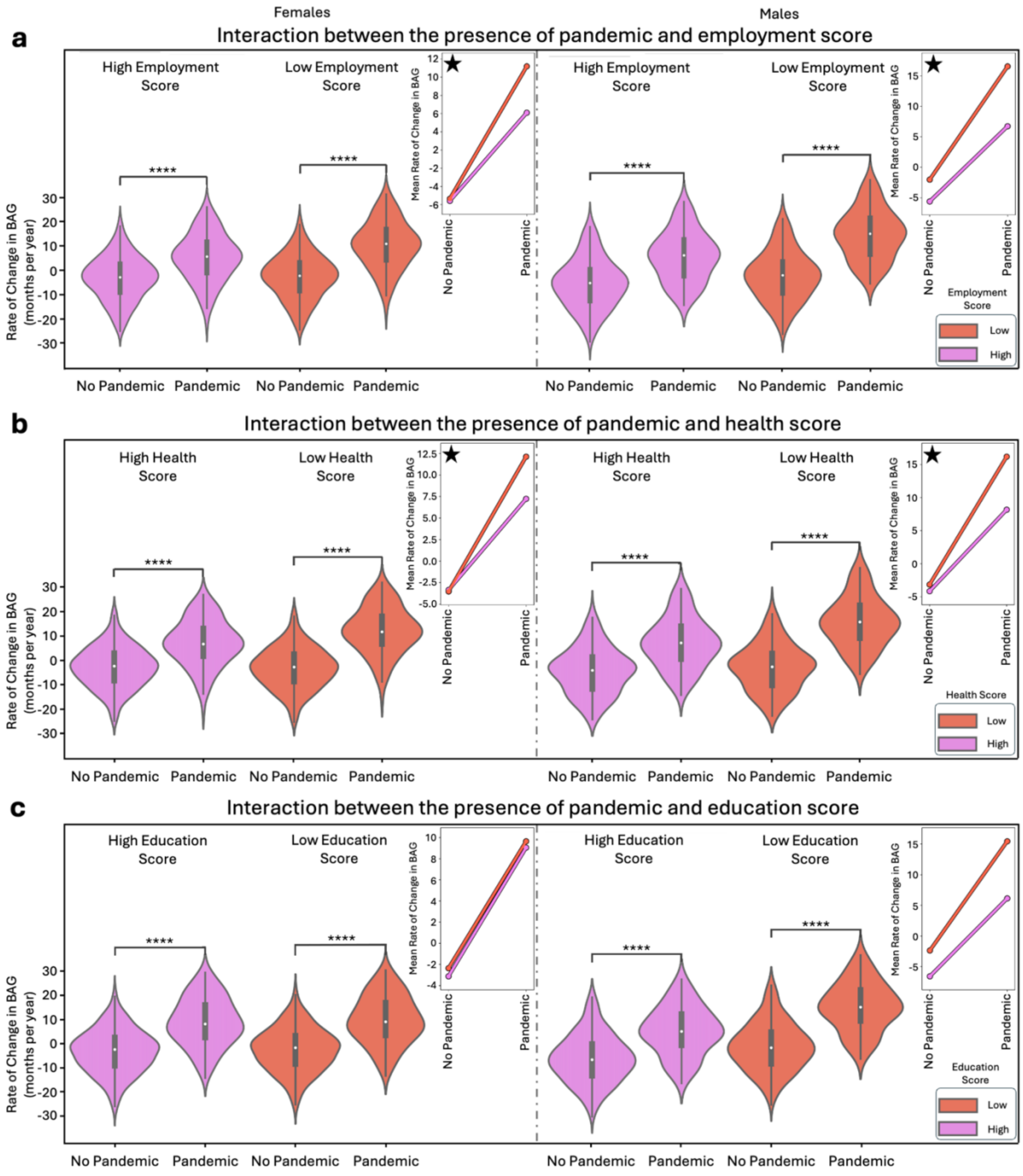


Fig. S5. Interaction of deprivation indices and pandemic status in grey matter models for female and male participants. The figure illustrates the distribution of the rate of change in brain age gap (BAG) for the "Pandemic" and "No Pandemic" groups within the grey matter (GM) model, analysed separately for female and male participants. The analysis is presented for three deprivation indices: (a) Employment score, comparing participants with low (purple) vs. high (red) levels. (b) Health score, comparing participants with low (purple) vs. high (red) levels. (c) Education score, comparing participants with low (purple) vs. high (red) levels. Each subplot contains two panels: the left panel displays the results for female participants, while the right panel shows the results for male participants. The findings indicate significant differences in brain ageing patterns between the "Pandemic" and "No Pandemic" groups across all deprivation indices, for both female and male participants in the GM model (p-values < 0.0001, indicated by **** for p-values ≤ 1.0e-04). Additionally, significant differences were observed between low and high deprivation groups (Employment score: females (p-value = 0.0160), males (p-value = 0.0002); Health score: females (p-value = 0.0188), males (p-value = 0.0022); Education score: significant in males only (p-value = 0.0002). Furthermore, significant interactions between pandemic status and deprivation factors were found (Employment score: females (p-value = 0.0258), males (p-value = 0.0258); Health score: females (p-value = 0.0130), males (p-value = 0.0104). All p-values are below the 95% confidence interval (CI) [0.0443–0.0564].

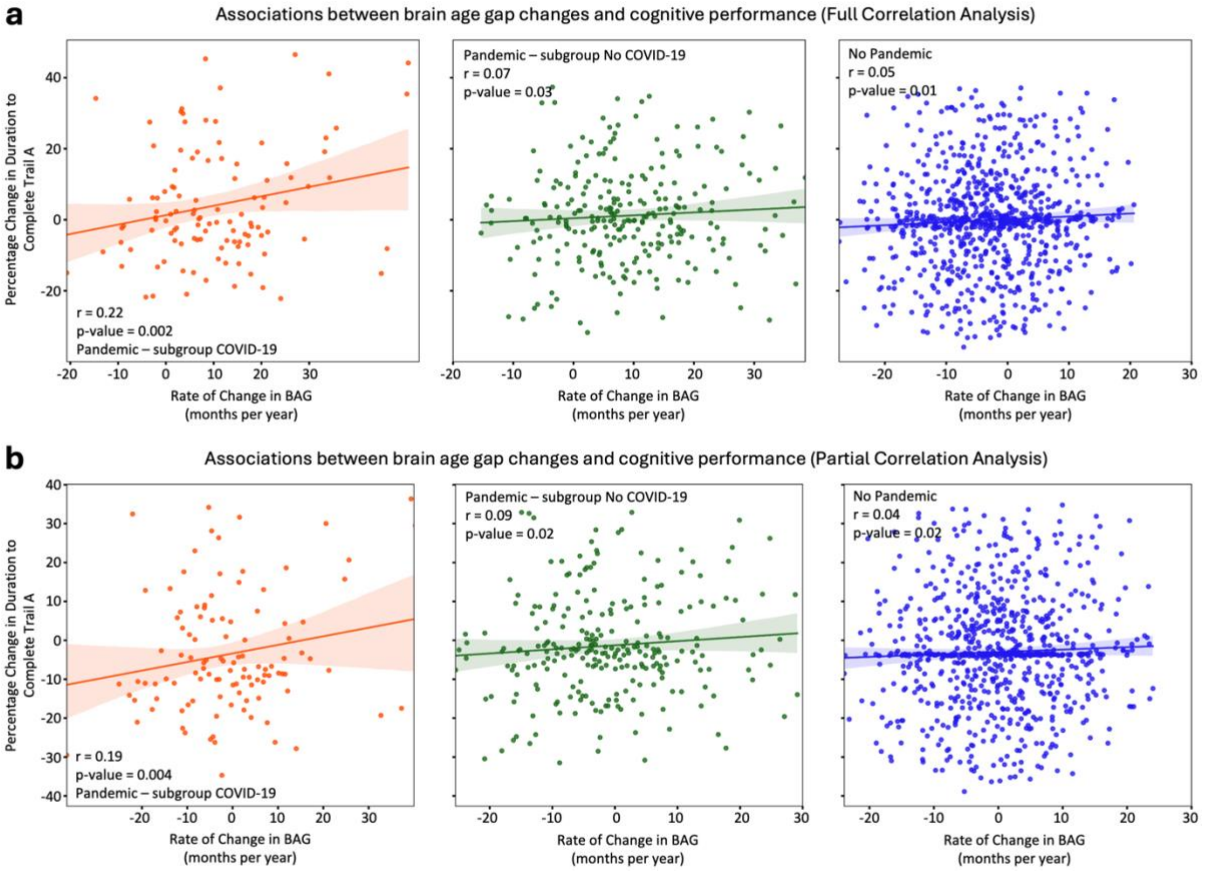


Fig. S6. The association between the rate of change in brain age gap and cognitive performance. The regression lines across different groups demonstrate a small but significant correlation between the rate of change in BAG and the percentage change in the duration to complete Trial A across different scans using (a) full correlation analysis and (b) partial correlation analysis, with the effect of age regressed out.
